# Interactions among 17 respiratory pathogens: a cross-sectional study using clinical and community surveillance data

**DOI:** 10.1101/2022.02.04.22270474

**Authors:** Roy Burstein, Benjamin M. Althouse, Amanda Adler, Adam Akullian, Elizabeth Brandstetter, Shari Cho, Anne Emanuels, Kairsten Fay, Luis Gamboa, Peter Han, Kristen Huden, Misja Ilcisin, Mandy Izzo, Michael L. Jackson, Ashley E. Kim, Louise Kimball, Kirsten Lacombe, Jover Lee, Jennifer K. Logue, Julia Rogers, Erin Chung, Thomas R. Sibley, Katrina Van Raay, Edward Wenger, Caitlin R. Wolf, Michael Boeckh, Helen Chu, Jeff Duchin, Mark Rieder, Jay Shendure, Lea M. Starita, Cecile Viboud, Trevor Bedford, Janet A. Englund, Michael Famulare, the Seattle Flu Study and SCAN Investigators

## Abstract

**Background:** Co-circulating respiratory pathogens can interfere with or promote each other, leading to important effects on disease epidemiology. Estimating the magnitude of pathogen-pathogen interactions from clinical specimens is challenging because sampling from symptomatic individuals can create biased estimates.

**Methods:** We conducted an observational, cross-sectional study using samples collected by the Seattle Flu Study between 11 November 2018 and 20 August 2021. Samples that tested positive via RT-qPCR for at least one of 17 potential respiratory pathogens were included in this study. Semi-quantitative cycle threshold (Ct) values were used to measure pathogen load. Differences in pathogen load between monoinfected and coinfected samples were assessed using linear regression adjusting for age, season, and recruitment channel.

**Results:** 21,686 samples were positive for at least one potential pathogen. Most prevalent were rhinovirus (33·5%), *Streptococcus pneumoniae* (*SPn*, 29·0%), SARS-CoV-2 (13.8%) and influenza A/H1N1 (9·6%). 140 potential pathogen pairs were included for analysis, and 56 (40%) pairs yielded significant Ct differences (p < 0.01) between monoinfected and co-infected samples. We observed no virus-virus pairs showing evidence of significant facilitating interactions, and found significant viral load decrease among 37 of 108 (34%) assessed pairs. Samples positive with *SPn* and a virus were consistently associated with increased *SPn* load.

**Conclusions:** Viral load data can be used to overcome sampling bias in studies of pathogen-pathogen interactions. When applied to respiratory pathogens, we found evidence of viral-*SPn* facilitation and several examples of viral-viral interference. Multipathogen surveillance is a cost-efficient data collection approach, with added clinical and epidemiological informational value over single-pathogen testing, but requires careful analysis to mitigate selection bias.

## Introduction

The human respiratory tract hosts a complex community of co-circulating pathogens and commensal bacteria. Competition for resources and modification of the host environment can act to inhibit or support pathogen replication, which can alter the natural history and severity of disease and ultimately impact population epidemiology.^1–6^ Furthermore, as multi-pathogen molecular testing becomes more common, increased understanding of coinfections will improve care at the bedside.

While literature on the interactions between pathogen pairs is mixed, viral-bacterial interactions are thought to be favorable for bacterial proliferation while viral-viral interactions are often considered inhibitory to replication. In particular, *Streptococcus pneumoniae* (SPn) colonization or increased proliferation following influenza infection is well documented,^7,8^ and evidence for similar facilitation following other respiratory viral infections exists.^9,10^ Pathogen interactions have typically been studied through laboratory experiments,^11–14^ animal models,^15–20^ and via transmission modeling,^5,21–25^ which have revealed important interspecific nuances within viral and bacterial pathogen relationships. While viral-viral interference is generally expected via innate immune responses, this generality does not always hold.^20^

Laboratory experiments studies are extremely useful but are laborious and costly. Epidemiological studies using multiplexed or arrayed polymerase chain reaction (PCR)-based techniques have explored pathogen-pathogen interactions based on their relative risk of co-occurrence; i.e., if two viruses co-occur less often than chance would predict, viral interference is concluded.^13,26,27^ However, such studies are prone to Berkson’s bias, a form of selection bias where under-representation of non-infected samples in clinical data will lead to spuriously low relative risks.^28–30^

Here, we develop a new approach for analyzing pathogen-pathogen interactions based on detecting differences among semi-quantitative measurements of pathogen load. This approach overcomes a number of shortcomings inherent to previous approaches based on binary co-occurrence in non-random samples by focusing on a within-subject measure of pathogen interaction, thus directly quantifying the direction and strength of interaction. We apply this method to a large sample of arrayed PCR data covering 17 potential pathogens or pathogen groupings from a large number of clinical samples collected by the Seattle Flu Study, a community-based respiratory surveillance platform.

## Methods

This study uses cross-sectional data collected through the Seattle Flu Study (SFS), including from the greater Seattle Coronavirus Assessment Network (SCAN). Participants enrolling in cross-sectional arms of SFS between 11 November, 2018 to 20 August, 2021 were eligible for this study. As previously described,^31,32^ nasal swab specimens were collected from individuals who self-reported at least two new or worsening symptoms of acute respiratory illness (ARI) or a new or worsening cough alone. Self-administered mid-turbinate or anterior nares swabs were collected at community sites including but not limited to transit stations, outpatient clinics, and at-home swab collection^33^. Residual specimens from patients tested for suspected respiratory infection at four regional hospitals were also collected. Only the first positive sample from each individual was used in this analysis. Participant age and date of swab collection were recorded for each specimen. Only samples tested for all included pathogens were retained, with the exception of SARS-CoV-2, which was not tested prior to 2020. **Supplementary Table 1** lists the recruitment sites used in this study. This study followed the STROBE and STROME-ID reporting guidelines.^34,35^

Each specimen was screened for a panel of potential respiratory pathogens in duplicate using a custom TaqMan OpenArray RT-PCR platform (Thermo Fisher).^32,36^ The following pathogen targets were included in this study: adenovirus (AdV); human coronaviruses (CoVs) 229E and OC43, HKu1 and NL63; human metapneumovirus (hMPV); human parainfluenza viruses (hPIV) 1 and 2, and hPIV 3 and 4; influenza A (IAV) H1N1 and H3N2; pan influenza B (IBV); pan influenza C (ICV); respiratory syncytial viruses (RSV) A and B; human rhinoviruses (RV); enterovirus D68 (EV.D68); pan enterovirus excluding D68 (EV); and *Streptococcus pneumoniae* (SPn). Specimens collected after 01-Jan-2020 were also tested for SARS-CoV-2 using a separate quantitative-PCR (qPCR) assay, previously described.^31^ SARS-CoV-2 Ct values were obtained by averaging two Ct values for the Orf1b gene primer. About 10% of SARS-CoV-2 samples were tested using a research-only OpenArray assay instead. We note that because of assay limitations, epidemiologically distinct strains were grouped into one assay each for CoV, hPIV, and RV. For conciseness, we refer to these pathogen groupings, and the potential pathogens AdV, RV, and SPn that may not be indicative of disease, simply as pathogens.

### Measurement of pathogen load

For pathogens tested by OpenArray (all except SARS-CoV-2), a relative threshold value (Crt) was computed for each positive result.^36–38^ Crts have a similar interpretation to qPCR cycle threshold (Ct) values as an inverse-proxy of viral or bacterial load in a sample, wherein lower values correspond to fewer cycles to reach a sufficient OpenArray signal, with each cycle roughly equivalent to a 2-fold reduction in genomic copies.^36,37^ For analysis of SARS-CoV-2, both Ct and Crt values were used, depending on the assay used (qPCR or OpenArray). We will refer to all cycle values generically as Ct.

### Statistical Analysis

For each pathogen-pathogen pair, we computed the average Ct difference between monoinfected and coinfected samples. For example, to test the effect of RSV-A on IAV-H1N1 infection, we compared the mean difference in H1N1 Ct values between samples that were only positive for H1N1 and samples that were positive for both H1N1 and RSV-A. Likewise, to test the effect of H1N1 on RSV-A (notated as H1N1→ RSV-A), we compared the Ct values between samples with only RSV-A detected and the RSV-A Cts for samples showing co-infection with H1N1. This was done for each pathogen-pathogen pair with sufficient sample size.

To control for potential confounding we tested Ct differences using linear regression, including age category (<1, 1-4, 5-17, 18-49, 50-64, 65+ years), calendar time (fixed effects for each year-month to account for non-linear trends), and recruitment mode category (community, outpatient clinic (kiosk), outpatient clinic (residual), and hospital residual) as control variables. For SARS-CoV-2, we also controlled for the assay used. Each pathogen comparison pair was run as an independent regression, and only samples either singly or coinfected were included. For example, to test the effect of RSV-A on H1N1 Ct, only samples that were either positive only for H1N1 or that were positive for RSV-A and H1N1 were included and H1N1 Ct values were used as the dependent variable:

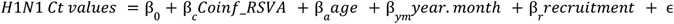

where *Coinf*_*RSVA* is a dummy variable indicating whether the sample was coinfected with RSV-A (0/1 for monoinfected/coinfected), and β *c* represents the average adjusted Ct value difference by which coinfected samples differ from monoinfected samples. Since Ct values are on an exponential scale and inverse to viral load, a coefficient of −1 represents approximately a two-fold average increase in genetic material in coinfected samples relative to monoinfected samples, suggesting a facilitating relationship. A coefficient of +4 represents an average of approximately a 2^4 = 16-fold decrease, suggesting interference. A p-value threshold of 0·01 was used for reporting significance to mitigate spurious significant findings across many pathogen pairs and we limited analysis to pairs with greater than 10 coinfections and 10 single infections.

We tested overall viral interaction effects similarly. For each pathogen, we tested the mean Ct interaction effect on all other viruses (ex. H1N1→ All viruses), as well as the mean effect of all viruses on each pathogen (ex. All viruses → H1N1). For the former, we included virus-specific fixed effects in each regression to control for baseline differences in Ct across viruses.

All analyses were performed using R version 4·1·2. Analysis code to be posted at https://github.com/InstituteforDiseaseModeling/SFS-coinfection-interactions

### Ethics approval statement

This study was approved by the University of Washington Institutional Review Board, with reliance from Fred Hutchinson and Seattle Children’s Hospital.

## Results

Of 115,087 samples collected and tested for multiple pathogens between 11 November, 2018 and 20 August, 2021, 32,894 (28·6%) were positive for at least one of the 17 pathogens included in this study. After exclusion of data from cohort sub-studies, multiple testing of individuals in cross-sectional data, and samples missing metadata, 21,686 positive samples and 27,422 total infection cases were retained for analysis. 11,182 (51·5% of total) positive samples were from hospital residuals, 7,633 (35·2%) from community testing (including 5720 from SCAN), and 2871 (13·2%) were from outpatient clinics. Among positive samples, the most prevalent pathogen was RV, present in 33·5% (N=7278) of samples, followed by *SPn*, a common commensal with the ability to cause disease, present in 29·0% (N=6293) of samples. Other common pathogens were SARS-CoV-2 (13·8%), IAV-H1N1 (9·6%), and IBV (7·8%). Of the positive samples, 16,692 (77·0%) had only one pathogen detected, while 4207 (19·4%) had two, 693 (3·2%) had three, and 94 (0·4%) had four or more. The most commonly co-occurring pathogen pair was *SPn* and RV, co-appearing in 1461 observations, followed by *SPn* and AdV (N=487), and *SPn* and RSV-A (N=423).

Average Ct values vary across pathogens, but in the absence of standardization, Ct values are not comparable across targets. *SPn* had the largest observed difference in Ct between monoinfected samples (20·7) and coinfected samples (17·9) (**Table 1**). Samples from hospitals and clinics generally had lower average Cts compared with community samples. For example, IAV-H1N1 community samples had an average Ct of 20·0, versus 16·3 from hospital samples. Ct was distributed differentially across ages by pathogen (**Figure 1**). Many pathogens, such as RSV-A, RSV-B, and IAV-H1N1 had lower average Ct values for children and elderly. Other pathogens, such as the seasonal coronaviruses, had flatter Ct distributions across ages. Children had higher prevalence of pathogen and coinfection detection than adults (**Supplementary figure 2**): of 6877 positive samples from children under 5 (representing 61·5% of our sample), 4732 (68·8%) were positive for multiple pathogens, while of 7177 samples from adults 18-49, only 836 (11·7%) were coinfections.

**Table 1:** Sample characteristics. **https://docs.google.com/spreadsheets/d/12C12sFn6QJdiMgq3aBavEukFf883unh9RDjR8kqWn5s/edit?usp=sharing**

|  | Adenovirus | Seasonal Coronavirus |  | Enterovirus |  | Metapneumovirus | Parainfluenza |  | Influenza A |  | Influenza B | Influenza C | Respiratory syncytial virus |  | Rhinovirus | Coronavirus (2019) | Strep. Pneumoniae |
| --- | --- | --- | --- | --- | --- | --- | --- | --- | --- | --- | --- | --- | --- | --- | --- | --- | --- |
|  | AdV | CoV 229E and OC43 | CoV HKU1 and NL63 | EV | EV D68 | hMPV | hPIV 1 and 2 | hPIV 3 and 4 | IAV H1N1 | IAV H3N2 | IBV | ICV | RSV A | RSV B | RV | SARS-CoV-2 | SPn |
| **Overall** |  |  |  |  |  |  |  |  |  |  |  |  |  |  |  |  |  |
| **All Samples** | 20.9 (sd: 6.3, N: 1484) | 16.4 (6.3, 1022) | 18 (5.5, 981) | 23.3 (3.2, 231) | 19.1 (6.3, 76) | 16.1 (5.5, 910) | 18.1 (5.5, 290) | 15.4 (6, 697) | 17.3 (5.2, 2074) | 15.6 (5.4, 896) | 14.1 (5.6, 1597) | 17.6 (6.8, 194) | 14.6 (5.7, 1079) | 15.6 (5.7, 632) | 18.9 (5.6, 7267) | 21.1 (7.7, 1699) | 19 (5.2, 6293) |
| **Coinfection status** |  |  |  |  |  |  |  |  |  |  |  |  |  |  |  |  |  |
| **Monoinfection** | 21.1 (6.2, 644) | 15.7 (6.1, 652) | 17.3 (5.4, 607) | 23.3 (3.2, 141) | 19.8 (6, 37) | 15.8 (5.3, 565) | 17.1 (5.4, 187) | 15 (5.9, 380) | 17.4 (5.3, 1570) | 15.5 (5.3, 674) | 14.1 (5.6, 1041) | 16.3 (6.7, 83) | 14.4 (5.6, 495) | 15.2 (5.5, 349) | 18.7 (5.7, 5217) | 21.2 (7.7, 1496) | 20.7 (4.8, 2554) |
| **Coinfection** | 20.8 (6.4, 840) | 17.5 (6.5, 370) | 19.2 (5.5, 374) | 23.2 (3.4, 90) | 18.5 (6.6, 39) | 16.5 (5.6, 345) | 20 (5.3, 103) | 15.8 (6.1, 317) | 16.8 (5.1, 504) | 15.7 (5.5, 222) | 14.1 (5.5, 556) | 18.5 (6.7, 111) | 14.6 (5.8, 584) | 16 (5.8, 283) | 19.3 (5.3, 2050) | 20.1 (7.7, 203) | 17.9 (5.2, 3739) |
| **Age** |  |  |  |  |  |  |  |  |  |  |  |  |  |  |  |  |  |
| **<1** | 19.1 (5.9, 229) | 14.5 (6.5, 85) | 18.1 (5.8, 82) | 23.3 (2.9, 28) | 26.9 (0.4, 4) | 15.1 (5.8, 89) | 14.5 (4.3, 19) | 14.8 (6, 98) | 13.6 (4.5, 92) | 13.5 (7.4, 9) | 12.7 (5.1, 71) | 17.4 (7.3, 22) | 12.7 (5, 244) | 12.1 (4.6, 106) | 18.2 (5.3, 689) | 16.4 (8.2, 54) | 17.4 (4.4, 714) |
| **1-4** | 20.3 (6.3, 596) | 15.5 (6.8, 199) | 18.3 (5.7, 196) | 22.8 (3.7, 82) | 21.7 (5.3, 7) | 15.3 (5.2, 270) | 17.9 (5.2, 113) | 14.5 (6, 278) | 15.7 (4.8, 270) | 15 (4.8, 85) | 13.8 (5, 296) | 18.4 (6.8, 81) | 13.8 (5.7, 428) | 15 (5.6, 189) | 17.9 (5.5, 1969) | 19.4 (7.8, 168) | 17.9 (5.1, 2358) |
| **5-17** | 20.5 (6.9, 220) | 17.5 (6.2, 119) | 17.5 (5.8, 140) | 22.8 (3.4, 49) | 19.6 (7.8, 10) | 16.2 (5.8, 123) | 18 (5.6, 62) | 15.9 (6.2, 101) | 15.6 (4.7, 470) | 14.7 (4.9, 192) | 13.1 (5.3, 662) | 18 (6.6, 31) | 16.1 (5.9, 139) | 17.3 (5.6, 49) | 18.6 (5.5, 1450) | 19.5 (7.6, 330) | 19.6 (5.2, 1259) |
| **18-49** | 22.9 (5.7, 320) | 16.5 (6, 368) | 18 (5.3, 376) | 23.7 (2.7, 39) | 18.7 (6.3, 37) | 17.4 (5.4, 227) | 19.9 (5.6, 57) | 17.4 (5.5, 119) | 18.4 (5.2, 748) | 15.9 (5.3, 320) | 15.4 (5.9, 480) | 14.9 (6.5, 40) | 17.3 (5.6, 130) | 17.8 (5.4, 122) | 19.5 (5.7, 2479) | 22 (7.6, 823) | 21.1 (5, 1249) |
| **50-64** | 22.4 (6, 77) | 17.2 (5.9, 151) | 17.7 (5.1, 127) | 24.5 (2.7, 20) | 17.9 (5.3, 13) | 16.4 (4.8, 103) | 18.4 (5.7, 22) | 15.2 (5.6, 53) | 18.9 (5.2, 325) | 17 (5.6, 148) | 16.1 (5.9, 58) | 19.4 (6.7, 15) | 15.9 (5.3, 79) | 16.9 (5.4, 91) | 20.2 (5.4, 443) | 21.6 (7.6, 200) | 19.7 (5.2, 498) |
| **65+** | 24.1 (4, 42) | 16.6 (6.2, 100) | 18.7 (5.9, 60) | 24.9 (1.9, 13) | 15.3 (5.8, 5) | 15.5 (5.6, 98) | 17.3 (6.1, 17) | 15.8 (6.2, 48) | 18 (5, 169) | 14.9 (5.8, 142) | 15.6 (5.9, 30) | 17.5 (5.3, 5) | 15.9 (6.1, 59) | 15.4 (5.5, 75) | 20.9 (5.7, 237) | 22.3 (7.2, 124) | 20.5 (5, 215) |
| **Recruitment** |  |  |  |  |  |  |  |  |  |  |  |  |  |  |  |  |  |
| **Clinic (Kiosk)** | 19.5 (6.9, 91) | 19.2 (6.4, 20) | 17.5 (5.8, 55) | 23.8 (2.8, 16) | 26.3 (--, 1) | 16.2 (4.8, 55) | 17.3 (5, 74) | 18.3 (6.2, 33) | 15.8 (5.3, 80) | 16.2 (6, 22) | 13.9 (5.6, 158) | 20.4 (7.3, 4) | 14.2 (5.1, 158) | 15 (6.1, 40) | 20.1 (4.2, 177) | -- | 18.1 (4.7, 348) |
| **Clinic (Flu VE Network)** | 21.9 (5.9, 55) | 17.1 (5.3, 227) | 17.3 (4.5, 76) | 22.7 (3.4, 28) | 18.4 (4.4, 6) | 16.5 (4.4, 150) | 19.3 (4.4, 35) | 16.3 (4.9, 100) | 19 (4.2, 348) | 16 (4.6, 441) | 16.8 (6.3, 13) | 17.1 (6.3, 34) | 14.4 (5, 152) | 15.7 (4.6, 154) | 18.5 (4.8, 223) | -- | 17.4 (3.6, 383) |
| **Community** | 23.7 (5.1, 386) | 17.3 (5.8, 163) | 18.2 (5.2, 295) | 24.7 (2.5, 22) | 20.3 (6.3, 26) | 18.6 (5.8, 92) | 19 (5.5, 48) | 15.9 (6.1, 69) | 20 (4.4, 309) | 16.9 (5.6, 31) | 17.6 (5, 250) | 19.3 (6.2, 5) | 18.5 (5.4, 82) | 17.7 (5.5, 54) | 18.6 (5.9, 3599) | 22 (7.4, 1389) | 20.9 (5.4, 1775) |
| **Hospital (Residual)** | 19.9 (6.4, 952) | 15.8 (6.6, 612) | 18.1 (5.8, 555) | 23.1 (3.3, 165) | 18.4 (6.5, 43) | 15.6 (5.6, 613) | 17.9 (6, 133) | 14.9 (6.1, 495) | 16.3 (5.3, 1337) | 14.9 (6, 402) | 13.3 (5.4, 1176) | 17.5 (6.9, 151) | 14.2 (5.9, 687) | 15.3 (6, 384) | 19.1 (5.4, 3268) | 16.9 (7.8, 310) | 18.4 (5.1, 3787) |

**Figure 1:**
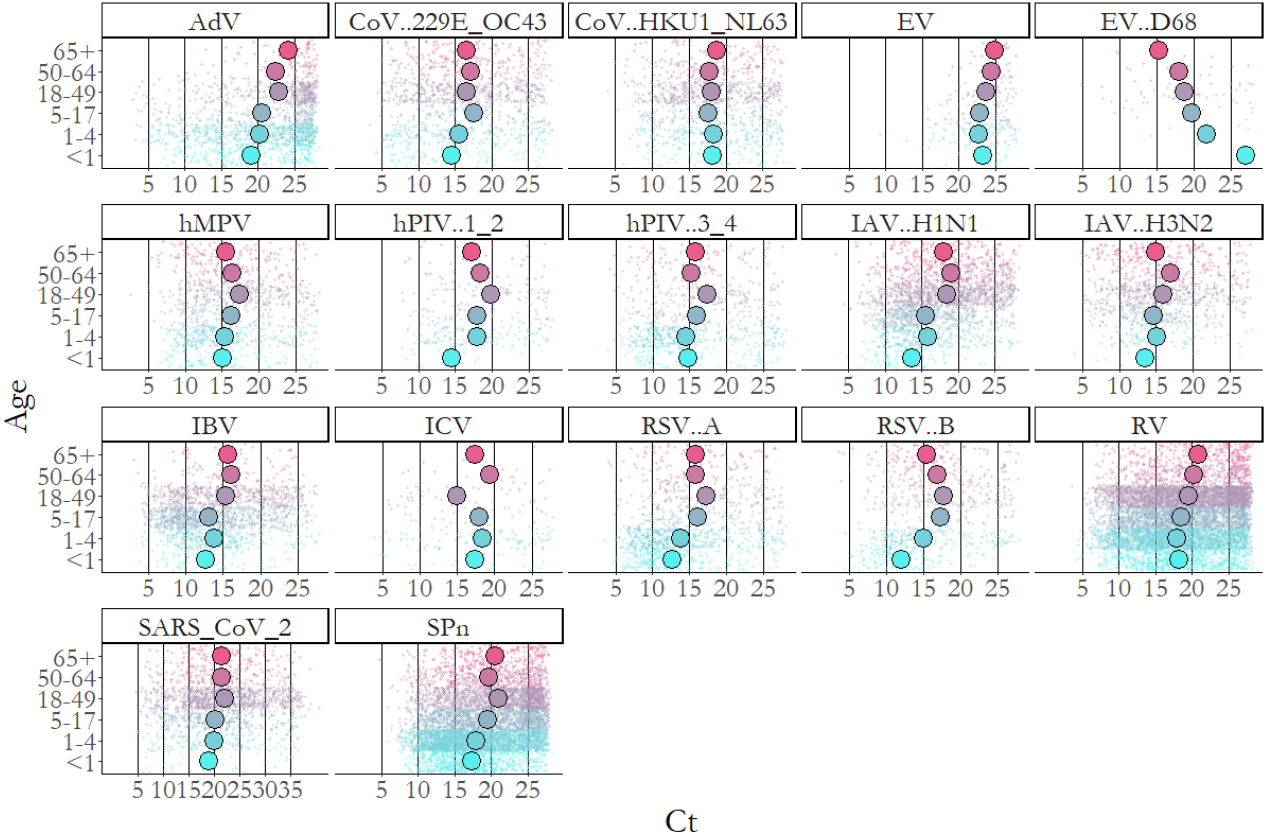
Ct distribution across age groups for each pathogen. Large dots represent mean values, each small dot represents the Ct value for one sample. Ct values range up to a maximum cycle of 28, except for SARS-CoV-2, for which the Taqman qPCR assay used ranged up to 40. Note that 146 (8·6% of total) SARS-CoV-2 Crt observations made using the OpenArray assay are not shown here to maintain consistency. We note that numerical Ct comparisons are meant to be within a target (across age) and not between targets.

Most samples come from the 2018-19 and 2019-20 winter seasons, when a large variety of respiratory pathogens were co-circulating (**Figure 2**). Each season had distinct characteristics. The 2018-19 influenza season was dominated by the A/H3N2 subtype, while the 2019-20 season had minimal circulation of A/H3N2 and was instead characterized by considerable IBV transmission. At the outset of the COVID-19 pandemic in March 2020, large-scale non-pharmaceutical interventions interrupted transmission of nearly all respiratory pathogens, with the exception of RV, *SPn*, AdV, and SARS-CoV-2, until February 2021, when other respiratory viruses began to reappear.

**Figure 2:**
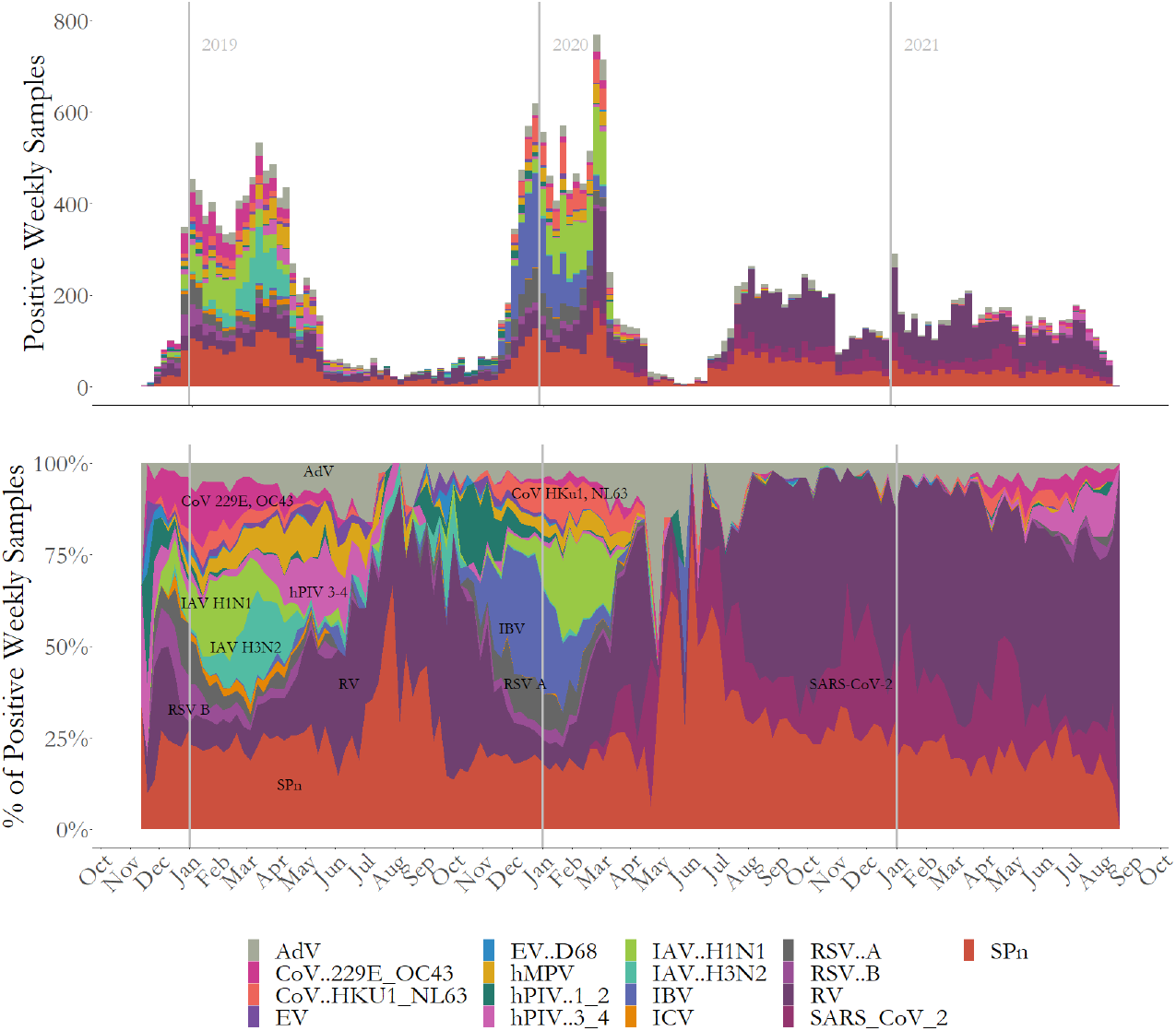
Total positive samples by week over the study period (top), and positive samples per week as a percentage of total weekly positive samples (bottom).

Of a total of 272 possible pathogen pairs for comparison, 132 had fewer than 10 coinfected samples, leaving a total of 140 pairs for testing. Of those, 56 (40%) pairs yielded absolute Ct differences that were significant at a 0.01 level (**Figure 3, Supplementary Data**). Ct differences ranged from −3 for (hMV and ICV) → *SPn* to +7 for IAV H1N1 → hMPV and (EV and IBV) → AdV. With the exception of *SPn*, there were no significant decreases in the Ct of any pathogen-pathogen pair. Samples positive with *SPn* and a virus were consistently significantly associated with decreased *SPn* Ct (in 12 of 16 viruses), with the exception of EV D68, EV, IAV H3N2,and hPIV 1-2. In contrast, Ct values for viruses were generally higher in samples coinfected with *SPn* than those without, significantly so for RV, IAV H1N1, RSV A, hMPV, hPIV 1 and 2, and all 4 seasonal CoVs.

**Figure 3:**
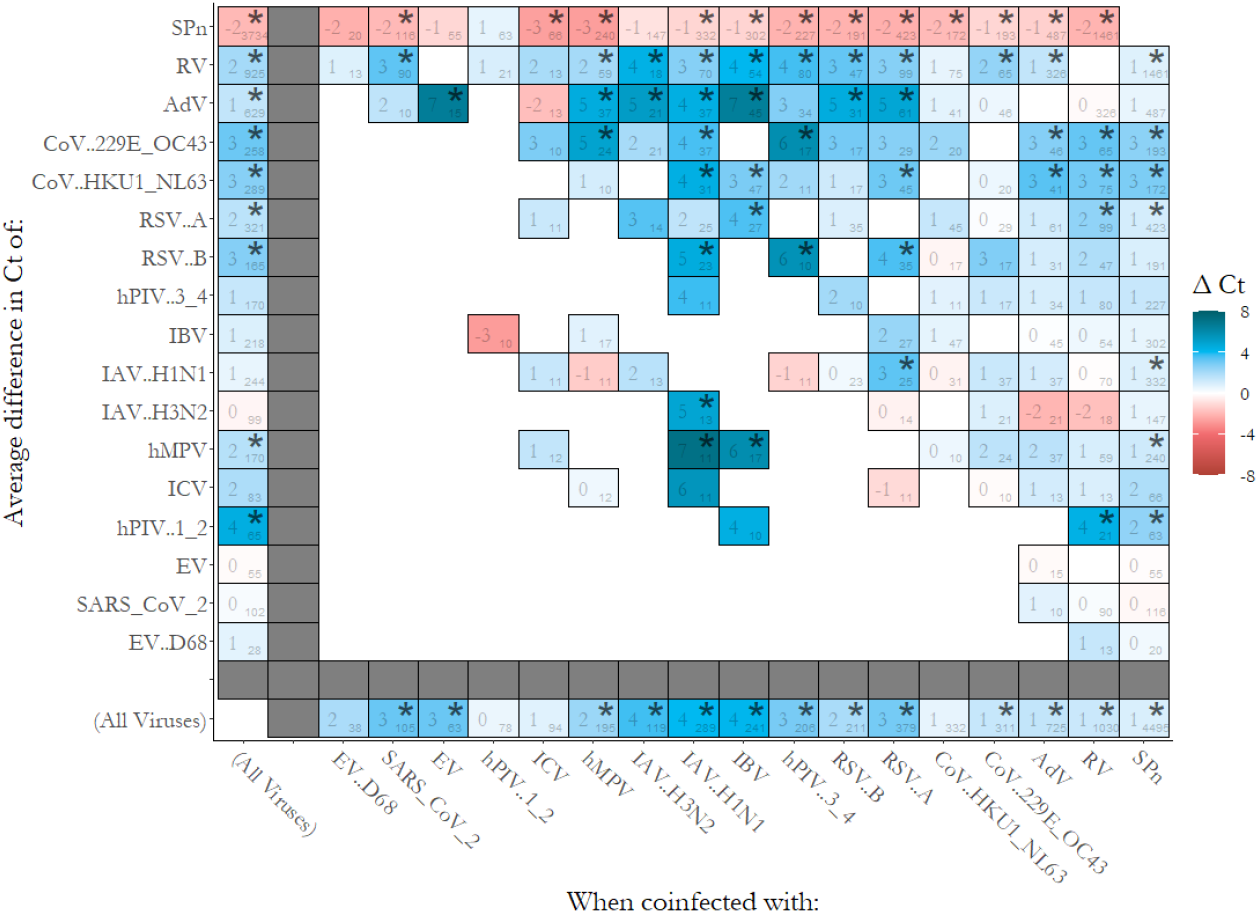
Adjusted Ct difference associated with each pathogen-pathogen pair, as well as average effects over all viruses (shown on margins). Colors reflect differences in Ct, with blues indicating an increase associated with interaction (suggesting interference), and reds indicating a decrease (suggesting facilitation). The large number in each square represents the average difference in Ct for mono-versus co-infections. The small number at the corner of each square represents the number of co-infected specimens in the sample. Stars (*) represent statistically significant Ct differences (p < 0·01). Pairs with fewer than 10 mono-or co-infected specimens were excluded. The first column and bottom row represent the average effects over all viruses. Full regression results summarized in this figure are available in the **Supplementary Data**.

We observed no virus-virus pairs that were significantly associated with decreased Ct during coinfection (which would be suggestive of facilitation), but found significant increases in Ct (suggestive of interference) among 37 of 108 (34%) of assessed pairs. Averaged viral interactions (shown on the left and bottom margins of Figure 3) reflected this trend as well. Coinfection with IAV and IBV was consistently associated with increased Ct in most other viruses (14 of 21 pairs), while coinfection with other viruses was associated with small or insignificant differences in IAV or IBV Ct values (2 of 21 pairs significant). The average virus had a +4 Ct value when coinfected with either IAV subtype or IBV, while we observed no significant change in either IAV or IBV when coinfected with other viruses, on average. Conversely, significant increases in the average Cts of CoVs were commonly observed when coinfected with other viruses (a +3 increase on average across all viral coinfections), but these viruses were not typically associated with large differences in the Cts of other viruses when coinfected (no individual significant interactions for the HKU1/NL63 target nor on average across all viruses, and 229E/OV43 only yielding a significant effect when coinfected with RV).

*SPn*, RV, and AdV were the only pathogens observed to co-occur in more than 10 samples with SARS-CoV-2, with 116, 90, and 10 co-occurrences, respectively. Of the individual six pairs tested, SARS-CoV-2 → *SPn* and SARS-CoV-2 → RV yielded significant results, with average Ct differences of −1·9 (suggestive of SARS-CoV-2 facilitating *SPn* replication) and +3·3 (suggestive of SARS-CoV-2 interfering with RV infection). Estimates of average viral interactions with SAR-CoV-2 are largely informed by RV and reflect this same trend: no change in SARS-CoV-2 Ct when coinfected, and a significant interfering effect on other viruses.

## Discussion

This study of 140 pairs of coinfections lends new empirical evidence on the dynamics of interspecies pathogen-pathogen interactions. We found no virus-virus pairs with evidence of facilitation -- where viral coinfection is associated with increased viral load relative to single infection -- but we found many suggestive of interference or competition -- where viral coinfection reduces viral load. In contrast, most viral→*SPn* coinfections were associated with significant increases in pneumococcal detection, suggestive of facilitation. These two broad findings generally conform with the existing body of evidence.^8^ Our results offer a wide-ranging survey that can serve to generate hypotheses for further research, particularly for pathogen pairs with little or no existing prior study.

Influenza A and B are the most potent suppressors of other viral infections in our study. Without exception, influenza coinfection reduced the viral load of co-infecting viruses, significantly so in 14 of 21 pairs with sufficient data to quantify. The effect was much weaker and rarer in the opposite direction. This finding suggests important directional distinctions which may not be independent of infection ordering and are likely mediated by virus-specific immune responses. For example, in a recent experiment in porcine tracheal epithelial cells, Peng and colleagues^14^ report 3-day prior influenza infection blocked coronavirus infection, while prior coronavirus infection had little effect on influenza replication. With the possible advent of more effective influenza vaccines, it will become important to monitor the impact on RSV, SARS-CoV-2, and other pathogens to mitigate possible unexpected shifts in overall burden and age distribution of other respiratory diseases.

A consistent increase in upper respiratory pneumococcal density associated with the presence of viral coinfectors has epidemiological and clinical implications. *SPn* is known to be commonly carried on the mucosal surface of the upper respiratory tract, but can cause severe disease when it becomes systemic and/or invades the lungs and other sterile sites.^39^ Innate immune responses to viral infections, particularly IAV, have been shown to increase bacterial adherence, colonization, and invasion, promoting disease and increasing *SPn* shedding, thus promoting onward transmission.^10,15,19,40–42^ Recent surveillance data from Israel shows that in the absence of circulating respiratory viruses during the COVID-19 pandemic, carriage remained relatively steady while pneumococcal disease declined.^43^ Likewise, we detected *SPn* in many samples throughout the COVID-19 pandemic (**Figure 2**), but found higher average density in samples with more viral coinfections at both individual (**Supplemental Figure 3**) and population levels (**Supplemental Figure 4**). Surprisingly, compared to most other viral coinfectors, the IAV → *SPn* relationship was among the weakest, suggesting potentially that other viral coinfectors may have a stronger facilitating relationships with *SPn*. This could be in part explained by our concurrent findings that IAV is associated with suppression of most other viruses (**Figure 1**), and that SPn replication is greater where more viruses are present (**Supplemental Figure 4**). Furthermore, and contrary to recent evidence,^44,45^ we did not observe a mutually facilitating relationship between IAV or any other virus and *SPn*.

Because of the dramatic effects of COVID-19 mitigation measures on all other respiratory pathogens, we only have sufficient sample sizes to characterize SARS-CoV-2 interactions between RV, AdV, and *SPn*. We found evidence to suggest that SARS-CoV-2 suppressed RV infection, with no additional evidence of interaction contrariwise. This finding contradicts recent studies which found an inhibiting effect of innate immune response to RV on SARS-CoV-2 infection.^46,47^ Among samples in our study, it is interesting to contrast our finding with the significant suppression of the hCoV family by RV coinfection. We speculate that prior immunity may be a deciding factor in the strength of RV:coronavirus competition. With only five endemic human coronaviruses compared with hundreds of endemic RV serotypes, the most common interaction may be coinfection with a RV serotype that the host is naive to and a coronavirus with some preexisting adaptive immunity. Prior adaptive immunity likely reduces peak viral load, and so we speculate that the ability of hCoV to interfere with RV is also lower, and the ability of RV to interfere with hCoV may be higher. This hypothesized effect of prior immunity for RV relative to SARS-CoV-2 could also explain the directional absence of impact from RV on SARS-CoV-2 observed in this study that contrasts with *in vivo* observations, and predicts that RV→SARS-CoV-2 interference may become more epidemiologically relevant in the future. Beyond the ecological interest in further studying this interaction, it may be informative about the role of population-level innate immune stimulation and/or within-host resource competition with other viruses for SARS-CoV-2 mitigation.^48,49^ Such competition has also been demonstrated between oral polio vaccine and influenza,^50^ and it thus may be an important generic concept for future pandemic suppression.

We designed our analysis to mitigate Berkson’s bias/collider bias ^28,51^ which affects all previous analyses of pathogen interaction based on multiple pathogen testing.^13,52–54^ Collider bias arises in this case due to studying symptomatic cases only, which enriches for positive samples.^29^ This reduces the number of pathogen-negative specimens in the sample relative to the general population, and yields biased inference on a 2×2 co-occurrence table that is sensitive to prevalence and pathogen-specific and pathogen-interaction effects on symptomaticity. While this issue has generally gone unrecognized in the literature, Nickbakhsh and colleagues^55^ explicitly attempted to mitigate it by limiting their analysis to virus-positive samples only. However, this meant that the ‘unexposed’ group was composed of samples positive for any virus other than the two of interest, yielding an analysis that is only interpretable relative to and highly sensitive to the composition of pathogens which were tested for and their prevalence. In contrast, our approach was based on comparing the density of genetic material from each pathogen when detected as monoinfections versus coinfections, and thus mitigating bias arising from under-inclusion of uninfected samples. While similar analyses have been conducted on smaller datasets,^40,56^ to the best of our knowledge, the present study is the most resolved and extensive yet in terms of specific pathogen-pathogen pair comparisons.

Like all studies, ours is subject to several limitations. First, for multiplex PCR assays that target more than one distinct pathogen, strain, or serotype (e.g. CoVs, AdV, RV, hPIVs, EV, *SPn*), our analysis could average over important heterogeneities in how each interacts with the other pathogens. RV is of particular concern as it comprises 3 species representing over 150 genotypes, with varying qPCR amplification efficiencies, resulting in inconsistent quantification of RV RNAs.^57^ As such, our results for RV could be sensitive to the genotypes circulating during this study. Second, several swab types were used over the study period without this data always collected and thus could not be directly adjusted in the analysis. We expect swab type leads to extra unexplained variation in Ct values but is unlikely to bias results as it is not associated with the exposure or outcome of interest. Third, infection and test timing, sequence, and inoculum size, are critical components of interaction dynamics,^14,58,59^ none of which we could observe or control for in this analysis; the lack of sequential sampling in our study also prohibits analysis of Ct changes in individuals over time. Future experimental, longitudinal, and modeling studies should be conducted to elucidate these factors. Fourth, while we included an extensive panel of 17 pathogens, it is possible that monoinfected individuals in our study are infected with unmeasured pathogens. Fifth, to the extent that pathogen load is correlated with symptomicity, we may be selecting for lower Ct values. This is a conservative bias, though, as it would affect both monoinfected and coinfected samples and would lead us to observe smaller differences in mean Ct.

Fifth, we cannot draw specific conclusions about transmissibility, disease severity, or symptom aetiology during coinfection as differences in Ct may correlate differently to relevant clinical parameters across pathogens and individuals.^60^ It is possible the association between aetiology and recruitment could act as another uncontrolled source of sampling bias. For example, X is more likely to lead to hospitalization than Y, Y may appear as a weaker ‘innocent bystander’ in coinfected samples; more experimental data is needed to fully understand these dynamics.

As multiplex PCR becomes cheaper, faster, and more available, its use as a clinical tool will increase dramatically. Future work should encompass understanding the clinical severity of specific pathogen-pathogen coinfections, which will be useful at the bedside for identifying high-risk combinations. With increasing ability to understand exactly the causes of infection, future vaccine development and implementation, treatments, and public health priorities could be tailored and thus reduce the extreme burden of respiratory infections affecting billions of individuals worldwide.

## Research in Context

### Evidence before this study

Evidence for interactions among respiratory pathogens has been typically limited to laboratory experiments involving animal or human cell models. These studies elucidate mechanisms but are typically limited in scope to one-on-one interactions. Multiplex PCR results from clinical datasets have been increasingly used to detect anomalies in the frequency of co-detection among pathogen pairs. Such studies benefit from larger sample sizes, and expanded scopes across many co-circulating pathogens, though relative risks of co-detection are biased in clinical samples due to Berkson’s bias, a form of selection bias where the uninfected are under-represented. We searched “viral co-occurrence” and “viral co-detection” and in PubMed and Google Scholar and only identified one paper which identified and attempted to address this selection bias.

### Added value of this study

To our knowledge, our study of respiratory pathogen interactions is the largest yet in terms of sample size and scope, with 21,686 cases across 17 pathogen groups. We developed an approach to detecting interactions that was not based on co-occurrence, but rather on pathogen load among positive samples, thus mitigating the effect of Berkson’s bias. Our study produced new statistical evidence for interactions among 140 pathogen pairs. We found no instances of increased viral load during viral-viral co-detection. Among other results, we found that influenza A and B were commonly associated with low viral load of co-infecting viruses, while there was little evidence that other viruses act to interfere with influenza.

### Implications of all the available evidence

Our study confirmed previous observations of viral infections leading to increased pneumococcal density, and broad viral-viral interference. The interspecific findings of our analyses add further nuance to these general understandings, to be further interpreted alongside experimental evidence. Past evidence generated from co-detection analyses may need to be reinvestigated as it is highly likely affected by selection bias. As multipathogen surveillance becomes ubiquitous, a richer understanding of coinfectin dynamics will be useful for clinical and epidemiological risk assessment.

## Data Availability

Data used for this study are are in the process of being prepared for open publication (as of 04-FEB-2022)

## Acknowledgements

We would like to thank the Seattle Flu Study and SCAN participants for their invaluable contributions to this research, and the entire Seattle Flu Study team for making this study possible. The Seattle Flu Study and SCAN are administered by the Brotman Baty Institute for Precision Medicine and funded by Gates Ventures, the private office of Bill Gates. JS is an Investigator of the Howard Hughes Medical Institute. RB and MF are employees of the Institute for Disease Modeling, a research group within, and solely funded by, the Bill and Melinda Gates Foundation. T.B. is supported by NIH R35 GM119774.

Ethics Approval: The Seattle Flu Study received approval by the University of Washington’s Institutional Review Board at the (UW IRB; STUDY00006181) and informed consent was obtained prior to study enrollment.

Participants participated in SCAN as part of public health

## Supplementary Information

- Table of regression results
- Table of sample sizes by recruitment site
- Supplementary Figures

1. Sensitivity analysis: Results if only single co-infections were included
2. Age and recruitment distributions of coinfections
3. Crt versus number of coinfectors by pathogen
4. *SPn* positivity versus number of viral coinfections in the population over time
5. Sample exclusions flow diagram

***Supplementary Data:*** *Full regression model results for each interaction summarized in Main Figure 3. Includes results for an unadjusted effect and the full model which adjusts for month, age, and recruitment mode*. ***https://docs.google.com/spreadsheets/d/1RfSaEnRhTuwQf5AEPB-blapupCr4DIlpCvvZBvwEskM/edit?usp=sharing***

**Supplementary Table 1:** Number of samples by recruitment site

| Recruitment Type | Site | Sample Size |
| --- | --- | --- |
| Clinic (Kiosk) | ChildrensHospitalSeattle | 621 |
|  | ChildrensHospitalSeattleOutpatientClinic | 168 |
|  | UWHallHealth | 43 |
|  | UWSeaMar | 40 |
|  | PioneerSquare | 27 |
|  | ChildrensHospitalBellevue | 25 |
|  | ChildrensSeaMar | 16 |
| Clinic (Flu VE Network) | KaiserPermanente | 1931 |
| Community | SCAN | 5720 |
|  | swabNSend | 1347 |
|  | WestlakeMall | 153 |
|  | HUB | 100 |
|  | UWSuzzalloLibrary | 83 |
|  | HarborviewLobby | 81 |
|  | FredHutchLobby | 69 |
|  | SeattleCenter | 12 |
|  | CapitolHillLightRailStation | 12 |
|  | Harborview | 10 |
|  | UWGreek | 10 |
|  | ColumbiaCenter | 6 |
|  | HealthSciencesRotunda | 5 |
|  | Costco | 5 |
|  | UWClub | 5 |
|  | KingStreetStation | 4 |
|  | SeaTacInternational | 3 |
|  | UWReopeningSwabNSend | 3 |
|  | **PICAWA** | 2 |
|  | **WestCampusChildCareCenter** | 1 |
|  | **SeaTacDomestic** | 1 |
|  | **UWOdegaardLibrary** | 1 |
| **Hospital (Residual)** | **RetrospectiveChildren'sHospitalSeattle** | 7600 |
|  | **RetrospectiveHarborview** | 2000 |
|  | **RetrospectiveNorthwest** | 982 |
|  | **RetrospectiveUWMedicalCenter** | 592 |

**Supplementary Figure 1:**
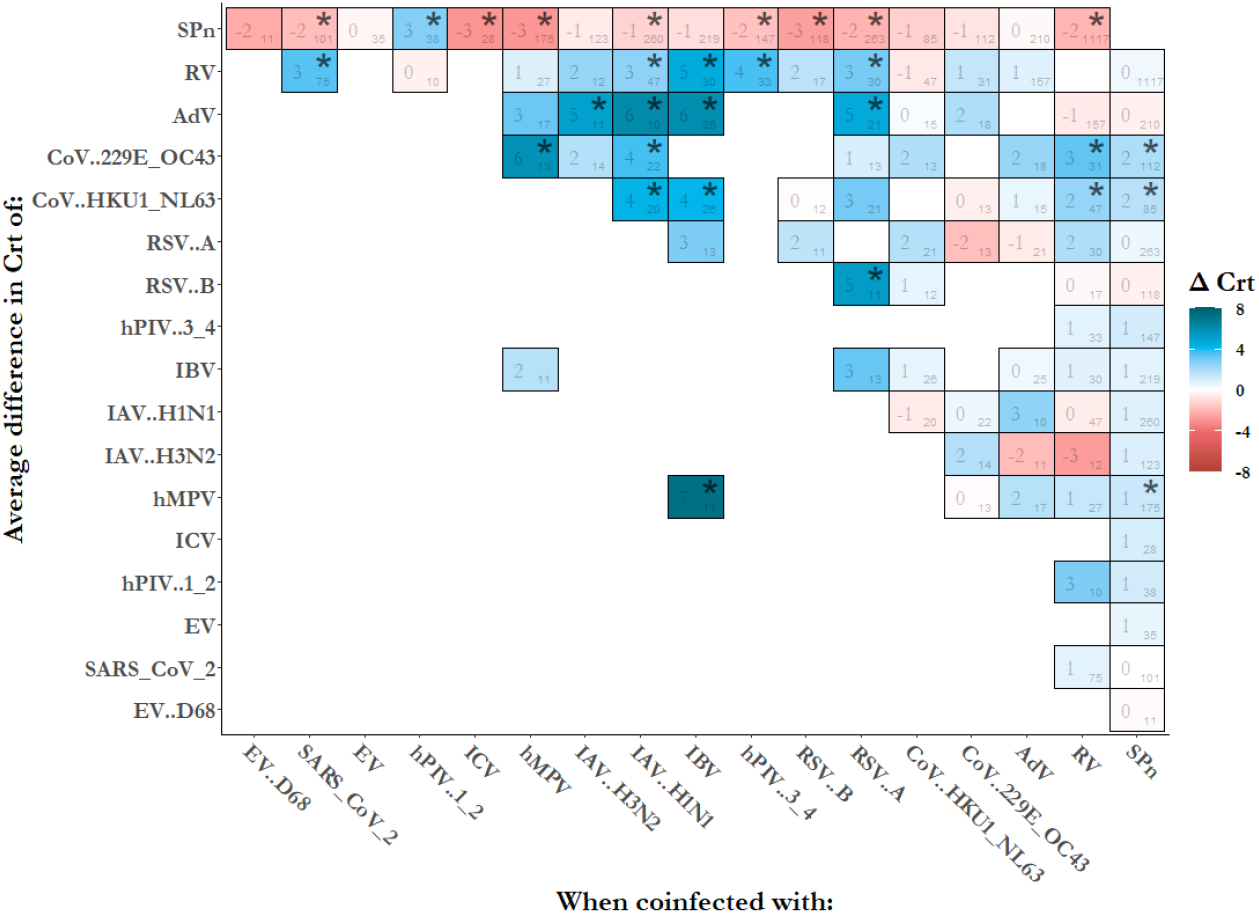
Sensitivity analyses - replication of main Figure 3. Keeping only monoinfected and coinfected samples with only two pathogens. Findings are largely qualitatively similar with some pathogen pairs removed as sample size is reduced.

**Supplementary Figure 2:**
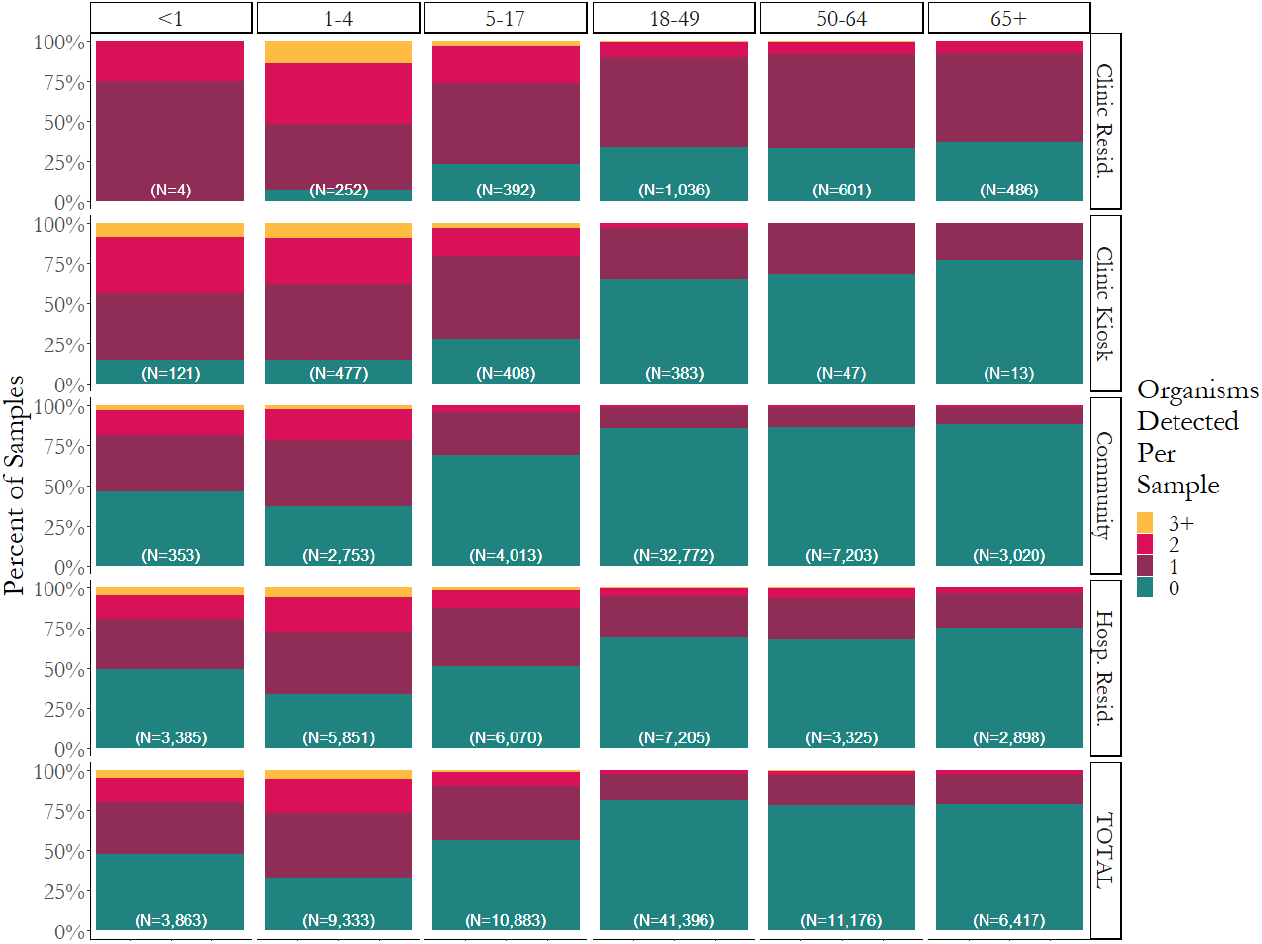
Frequency of sample infections by age and recruitment type across 83,068 samples with complete metadata. Children and adolescents are more likely than adults to have one or more infections.

**Supplementary Figure 3:**
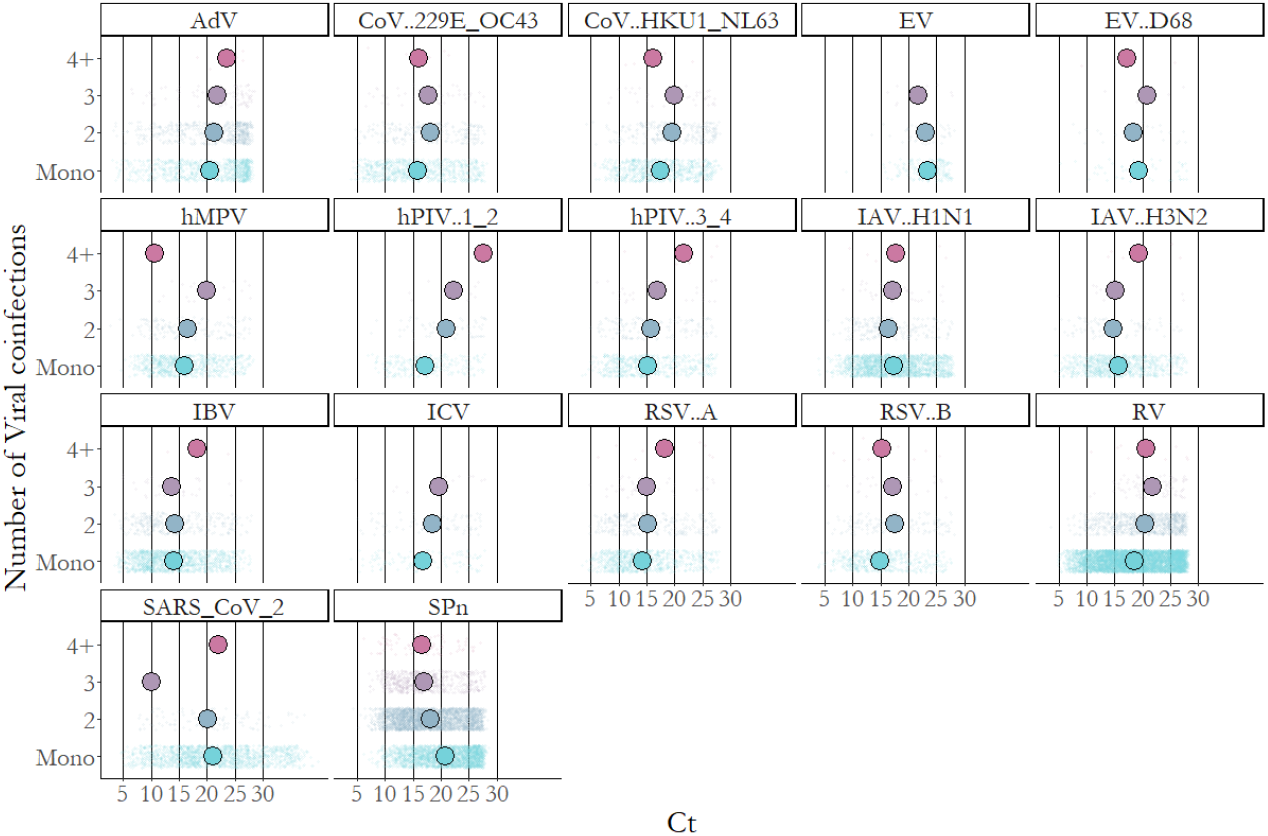
Average Ct values by number of viral coinfections. Strong gradient observed for SPn, where more viral coinfectors are strongly associated with more bacterial replication in the upper respiratory tract.

**Supplementary Figure 4:**
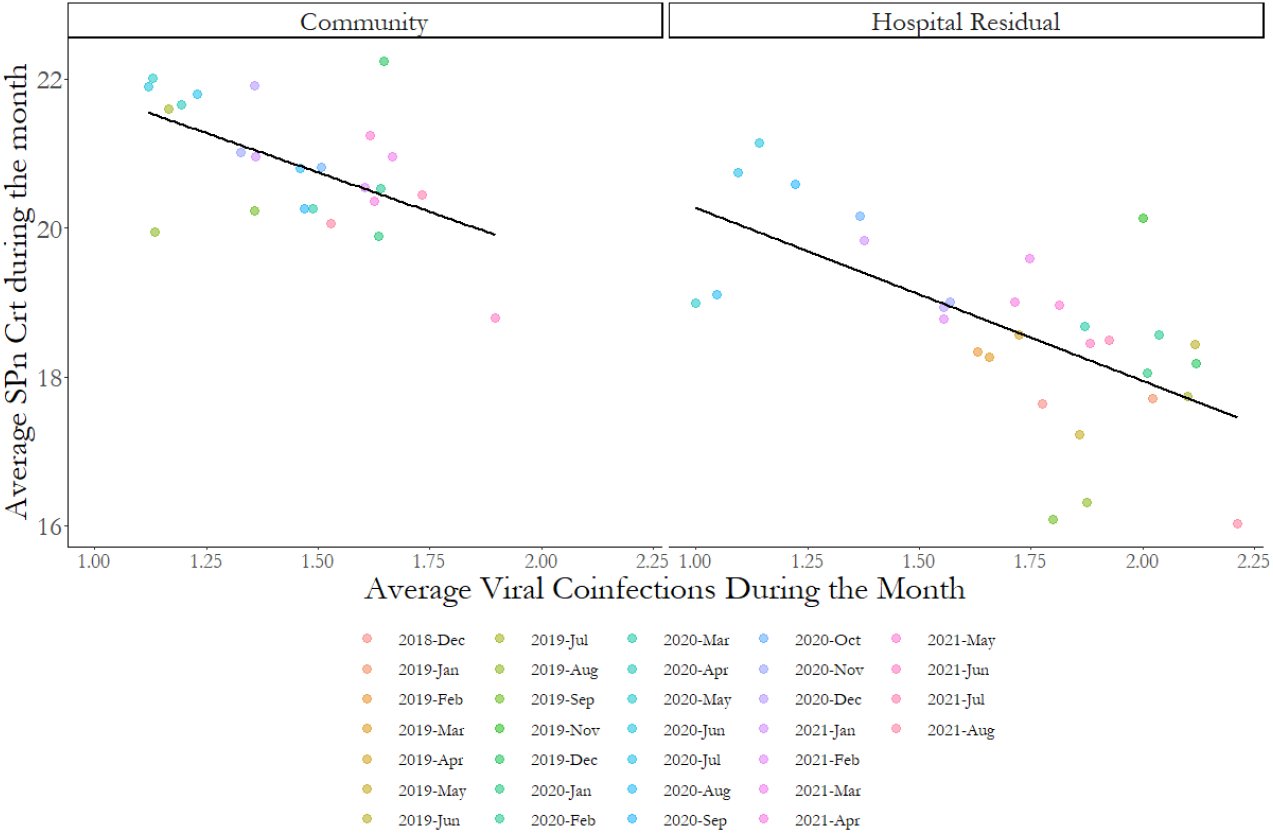
Ecological association between average number of circulating viruses in a month and the average Crt values for *SPn* in community and hospital samples, suggesting population-level evidence for the individual-level interaction explored in this paper.

**Supplementary Figure 5:**
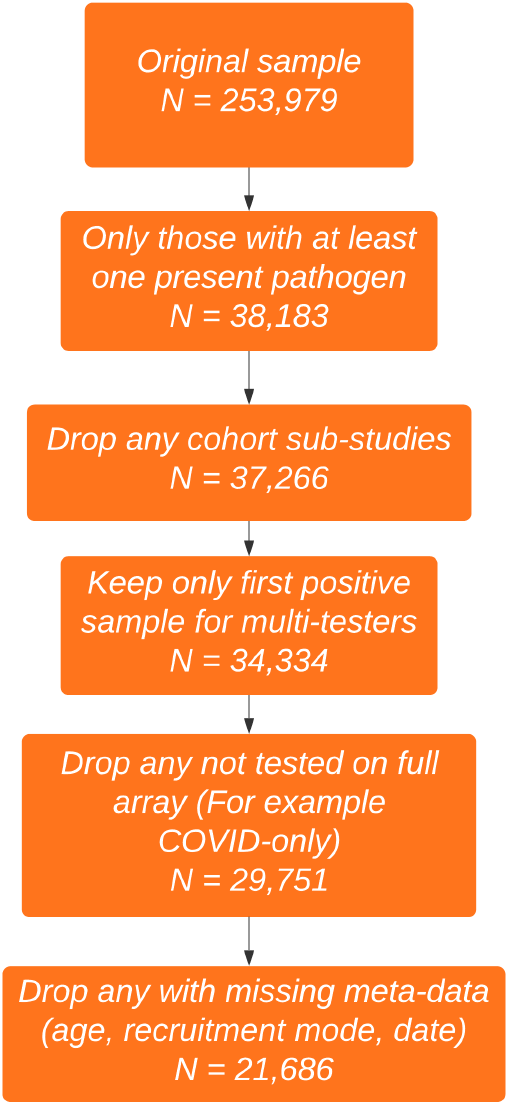
Sample exclusions

